# Self-Care from Anywhere: Evaluating the usability of an AI-powered HIV toolkit among adolescent girls and young women and healthcare providers in South Africa

**DOI:** 10.64898/2026.04.01.26349925

**Authors:** Simamkele Bokolo, Caroline Govathson, Laura Rossouw, Sinethemba Madlala, Sasha Frade, Shawna Cooper, Sarah Morris, Sophie Pascoe, Lawrence Long, Candice Chetty-Makkan

## Abstract

**Background:** HIV remains a major public health challenge in South Africa, with gaps in early diagnosis and linkage to care driving onward transmission. Adolescent girls and young women face barriers to timely care, including stigma, privacy concerns, and limited clinic access, while healthcare providers work in resource-constrained settings with high client volumes. We evaluated the Self-Care from Anywhere (SCFA) toolkit, an AI-enabled intervention comprising an AI Companion for AGYW and a provider-facing Clinical Portal to support HIV prevention, testing, and linkage to care. The AI Companion is designed to complement and extend human-delivered services, particularly in resource constrained settings, rather than replace in-person counselling.

**Methods:** We conducted an exploratory study to assess the usability, feasibility, and acceptability of the SCFA toolkit in Gauteng Province (November 2024-May 2025). AGYW engaged with the AI Companion, and a subset completed a simulated HIV self-testing activity with AI-delivered counselling. Pre and post-intervention surveys, including the System Usability Scale (SUS), were administered. Usability testing of the Clinical Portal involved healthcare providers using the toolkit without formal training to capture first impressions. A subset of AGYW and healthcare providers participated in separate focus group discussions or in-depth interviews. Quantitative data were analysed using descriptive statistics, and qualitative data were analysed thematically.

**Results:** A total of 97 AGYW were enrolled; 75.3% had completed high school and 91.8% were unemployed or full time students. Most participants (85.6%) self-reported HIV-negative status, and 63.9% reported sexual activity in the past 12 months. The AI Companion demonstrated high usability (mean SUS 87.7, SD 12.7) and was perceived as acceptable and useful, particularly for its personalisation and confidentiality features. Healthcare providers had a mean age of 34 years (SD 6.5), with about half serving as HIV testing and screening counsellors. Most providers rated the Clinical Portal’s ease of use, comprehension, and client support as positive to very positive, though 23% expressed concerns regarding workflow efficiency and their ability to manage additional client volume. Providers also highlighted the Clinical Portal’s value for case management.

**Conclusion:** AI-powered digital health tools, such as the SCFA toolkit, show potential to enhance user engagement and support care delivery, with high usability and acceptability demonstrated among AGYW and healthcare providers. Continued user-centred refinement is essential to ensure these tools remain responsive to the evolving needs and care contexts of diverse user groups.

## INTRODUCTION

HIV remains a major public health challenge in South Africa. Although testing and treatment services are widely available, persistent gaps in early diagnosis and linkage to care mean undiagnosed infections continue to drive onward transmission.^1–2^ For many, especially adolescent girls and young women (AGYW), stigma, privacy concerns, and the inconvenience of accessing healthcare facilities are significant barriers to timely testing and treatment.^3–5^ Long travel distances, limited clinic hours, and fear of discrimination further discourage engagement in care.^6^ Similarly, healthcare providers face parallel challenges of staff shortages, high client loads, and limited resources, limiting the time available for counselling and follow-up. These challenges undermine the quality of care and contribute to client dissatisfaction and disengagement from healthcare.^7–9^

Traditional models of healthcare, centered on clinic visits and in-person consultations, often fall short in addressing the specific challenges that adolescent girls and young women face.^3–4,10^ Digital health technologies, particularly artificial intelligence (AI), present an opportunity to complement these approaches through confidential, adaptive, and accessible platforms that support individuals outside of clinical settings.^11–12^ AI can help relieve the burden on overstretched providers by streamlining client management, triaging cases, and enabling efficient use of limited resources.^13–14^

Across global health systems, AI-enabled tools have shown the potential to enhance early diagnosis, support treatment adherence, and facilitate remote monitoring.^15–16^ In resource-limited countries, these tools may extend access to accurate health information, provide contextually sensitive support, and strengthen provider capacity.^17^ In HIV care, where stigma and human resource shortages remain critical challenges, AI may help deliver equitable and responsive services. With the expanding HIV prevention landscape, including daily oral and long acting injectable pre-exposure prophylaxis (PrEP), digital tools are an important avenue for education, demand creation, and linkage to prevention services. Understanding how young women use and experience these tools is therefore important.

AI tools such as conversational agents, including foundation model systems like ChatGPT, messaging based chatbots, mental health support bots such as Woebot and Wysa, and emerging computer-vision health applications are increasingly used to support personalised digital health guidance.^18–20^ Despite this growing ecosystem, most available tools are designed for broader consumer use and seldom reflect the unique realities of adolescents and young people seeking sexual and reproductive health or HIV-related counselling. Existing digital health tools often rely on generic scripts, provide limited emotional resonance and offer fragmented support that does not integrate counselling, self-testing guidance, decision making and linkage to clinical care.

Building on the potential of digital tools in healthcare and addressing limitations of existing approaches, the Self-Care from Anywhere (SCFA) toolkit developed by Audere [https://www.auderenow.org/] integrates multiple types of artificial intelligence into HIV prevention and treatment pathways to support sexual health care delivery. The toolkit comprises two interconnected user-facing components: (1) an AI Companion for clients, delivered via WhatsApp, designed to answer any health or lifestyle question, provide confidential guidance on HIV self-testing, empathetic pre- and post-test counselling, clear information on next steps with a private channel for engagement with healthcare providers; and (2) a Clinical Portal for healthcare providers. The Clinical Portal enables providers to remotely review and manage client interactions by presenting structured summaries of client/AI Companion conversations, computer vision-enabled interpretation of HIV self-test results, and AI-generated indicators such as predictive HIV vulnerability scores to support clinical prioritisation and anonymous follow-up. This study assessed the usability, acceptability, and feasibility of the SCFA toolkit by examining AGYW engagement with the AI Companion and healthcare providers’ experiences using the Clinical Portal.

## METHODS

### Study context and design

This exploratory mixed-methods study was conducted in Gauteng Province, South Africa, between November 2024 and May 2025. We collected survey data and participants took part in focus group discussions (FGDs) and in-depth interviews (IDIs). Gauteng is the most populous province in South Africa, with over 15 million residents, which constitutes about 26.3% of the nation’s total population and over 1.1 million adolescent girls and young women aged 15-24 years, with most residing in Gauteng.^21;22^ In 2022, Gauteng had an HIV prevalence of approximately 12%, the third lowest prevalence compared to other provinces, however had the highest number of AGYW living with HIV.^22–23^ Gauteng has several socio-economic challenges, such as high unemployment rates (39%), that make it a critical area of focus for addressing persistent issues in HIV prevention and treatment efforts.

### Study population

The target population for this study were young women aged 16-24 years, given their risk of HIV acquisition and challenges with accessing healthcare services. Healthcare providers enrolled in the study were involved in HIV prevention and treatment services from either the private or public sector.

#### Adolescent girls and young women

We invited AGYW through the [Shout-It-Now](https://shoutitnow.org/) programme, an implementing organisation that delivers HIV and sexual and reproductive (SRH) services to young people, historically targeting young women. Recruitment took place in the Tshwane and Ekurhuleni districts. We aimed to enrol 100 participants; 18-24 year olds (n=90) and 16-17 year olds (n=10). The larger target for adult young women was aligned with the age distribution of the population routinely reached through Shout-It-Now’s service delivery and reflects the typical age of HIV acquisition in this population.^24–25^ However, given the high HIV vulnerability among adolescent girls and the ability to reach them before the typical age of HIV acquisition in South Africa, we included 16-17-year-olds in the study. AGYWs were included if they self-identified as females, were 16-24 years, willing and able to provide written informed consent or assent, willing to self-report their HIV status at enrolment and were able to read and understand English.

#### Healthcare providers

We aimed to enrol 50 healthcare providers from the private and public healthcare sectors from Shout-It-Now and Indlela’s Behavioural Hub. The behavioural hub is a panel comprising healthcare providers and community members who have consented to participate in research studies.^26^ Providers were included in the study if they: 1) were ≥18 years, 2) understood English, 3) were comfortable providing consent, and 4) provided HIV prevention and treatment services. Providers who took part in the FGDs or IDIs were selected using convenience sampling, based on their availability to participate. This approach was necessary given the demanding schedules of healthcare providers.

### Sample size

The sample was not powered for statistical comparisons or hypothesis testing. However, our approach aligned with usability testing guidance, which supports the use of smaller, purposively selected samples to allow for in-depth exploration of user experience, rapid identification of usability issues, and iterative refinement of digital tools.^27^ Smaller samples are often sufficient to uncover the majority of usability problems, particularly when combined with qualitative methods such as focus group discussions and in-depth interviews.

### Study procedures

#### Adolescent girls and young women

Before data collection, written informed consent was obtained from those ≥18 years, including adolescent assent and parental consent from those <18 years. At enrolment, participants were assigned to one of the two study groups based on their self-reported HIV status: **Group A -** AI Companion interaction only or **Group B -** AI Companion interaction and a simulated HIV self-test (HIVST). Those who self-reported as HIV-negative or with an unknown status were assigned to Group A or Group B. Participants who reported living with HIV were assigned to Group A to avoid personal discomfort and confusion with the randomly allocated mock test results that formed part of the simulated HIVST in Group B.

Participants then completed a baseline (enrolment) survey that captured demographic information and assessed their intention to engage in HIV care services. Validated Likert-scale items were used to measure intention to seek HIV care at enrolment. Participants then completed the Balloon Analogue Risk Task (BART) in the Indlela Behavioural Lab to assess risk-taking propensity and to assess the relationship between risk taking propensity and willingness to engage with AI, results from the BART are reported elsewhere.^28^ After the BART, all participants interacted with the AI Companion, which provided tailored guidance on sexual health, relationships, and HIV prevention.

Those in Group B participated in a simulated HIV self-testing activity using one of three commercially available HIV self-test kits: Abbott CheckNOW, OraQuick HIV Self-Test and Wondfo HIV Self-test. Group B participants followed digital instructions provided on the SCFA toolkit, which provided step-by-step or summarised interactive guidance through the AI Companion describing how to perform the HIV self-test correctly. Participants received digital standardised post-test counselling corresponding to their assigned mock HIV test result (positive, negative, or invalid). For both groups, interactions with the AI Companion were observed by a study team member who documented navigation challenges and user experiences.

After completing the assigned activities, all participants completed a post-survey identical to the baseline to measure changes in their intention to engage in HIV care. The post-survey also included the System Usability Scale (SUS) and additional items assessing, acceptability, and satisfaction with the AI Companion.^29^

Finally, a subset of participants took part in focus group discussions (FGDs) to further explore the feasibility, acceptability, and satisfaction of interacting with the AI Companion and, where applicable, the simulated HIV self-test. Participants were purposively selected based on group assignment (Group A or Group B) and stratified age eligibility (16-17 years or 18-24 years), and consented to participate. FGDs were stratified by age group (16-17 years and ≥18 years) and by study group (Group A or B), with separate sessions for participants who only engaged with the AI Companion and those who also completed the simulated HIV self-test. Additional consent was obtained for participation and audio recording of the FGDs. The FGDs lasted 45-60 minutes and were co-facilitated by two female research assistants to make participants feel more comfortable and encourage open discussion. An FGD guide was used to facilitate the discussion (Refer to Appendix 1: FGD guide for AGYW).

#### Healthcare providers

Healthcare providers participated in a separate set of activities to explore their perceptions of the Clinical Portal. They provided feedback on the Clinical Portal’s capacity to support clinically appropriate, personalised care for clients.

Written informed consent was obtained, with additional consent for focus group discussions (FGDs) or in-depth interviews (IDIs) and for audio recording where applicable. Following consent, providers completed a demographic survey capturing personal details, highest qualification, profession, and years of professional experience (refer to Appendix 2: FGD/IDI guide for providers). Providers then watched two brief introductory videos that oriented them to the Clinical Portal’s features and capabilities. The videos served only to provide introductory familiarity with the Clinical Portal and were not meant to substitute for the full, structured training that healthcare providers ordinarily receive when new products or systems are implemented.

During the sessions, providers managed simulated client cases within the portal to evaluate its functionality and user-friendliness. The study team used a structured observation log to guide providers through the process and to record navigation performance and other observations.

Following this session, providers completed a post-survey assessing the effectiveness, ease of use, and satisfaction with the Clinical Portal for client management. A subset of providers participated in FGDs or IDIs to explore their experiences, focusing on ease of navigation, clarity of information, efficiency in completing tasks, satisfaction with integrated AI technologies, and willingness to adopt the tool in daily workflows.

### Data analysis

#### Quantitative analysis

Statistical analyses were performed using STATA version 18 (StataCorp, 2023). We used descriptive statistics to summarise participants’ socio-demographic characteristics and survey outcomes, reporting means and standard deviations (SD) for continuous variables and percentages for categorical variables. Descriptive statistics are reported separately for AGYW (client users) and healthcare providers. SUS scores were calculated and reported disaggregated by baseline characteristics (age group and sexual activity) and baseline engagement with HIV care (self-reported HIV and PrEP status, with PrEP referring to pre-exposure prophylaxis). Group differences were examined using 95% confidence intervals.

We report the six dimensions of the Intention to Engage in HIV care separately, comparing pre-and post-intervention average Likert Scores using confidence interval plots. It is important to note that dimensions five and six had six Likert Categories as opposed to five Likert categories, so the average scores could range from 1-5 (for dimensions 1 to 4) and 1-6 (for dimensions 5 and 6). Finally, healthcare providers’ perceptions of the Clinical Portal are presented in a heat map, with colour intensity indicating the concentration of responses across Likert categories.

#### Qualitative analysis

Qualitative data were analysed using NVivo version 12. Audio recordings were translated and transcribed. Thematic analysis with deductive and inductive approaches were used to analyse the data. Codebooks were developed and refined through an iterative approach by four researchers (SB, SM (HE^2^RO), CCM and a study team member). During discussions, inter-rater reliability across themes was assessed, where similar themes were retained while others were dropped when no consensus was reached. The study results were aligned with the User-Centred Design (UCD) framework.

### Ethics approval and consent to participate

The study protocol was approved by the University of Witwatersrand HREC Medical (ref: 240906) and Boston University Institutional Review Board (H-45383). Written informed consent was obtained from all participants prior to participation in the study and separate consent was obtained for FGDs and IDIs. The trial was registered with the South African National Clinical Trials Registry (https://sanctr.samrc.ac.za/), registration number: DOH-27-112024-5248.

## RESULTS

### Quantitative

#### Adolescent girls and young women

We initially aimed to enrol 100 AGYW. However, three participants cancelled shortly before data collection, resulting in a slightly smaller final sample. We enrolled 97 AGYW, of whom 41 (42%) participated in a simulated HIV self-test activity. The mean age of AGYW was 21 years (SD: 2.09), 75% completed high school education, 81% self-reported an HIV negative status and 63% had sex in the past 12 months. See Table 1.

**Table 1:** AGYW characteristics, SUS Scores by baseline characteristics and engagement with HIV care.

| **Age group** | **Outcome Measures** |  |  |
| --- | --- | --- | --- |
|  | **N(%)** | **SUS score** | **95% CI** |
| 16-17 years | 7 (7.2%) | 83.6 | 70.9-96.2% |
| 18-24 years | 90 (92.8%) | 88.1 |  |

|  |  |  | 85.5-90.6% |
| --- | --- | --- | --- |
| High school education |  | SUS score | 95% CI |
| Grade 8-11 | 24 (24.7%) | 80.6 | 74.7-86.5% |
| Grade 12 (completed high school) | 73 (75.3%) | 90.1 | 87.5-92.6% |
| Highest tertiary qualification obtained |  |  |  |
| Certificate | 12 (12.4%) | 89.6 | 83.3-95.9% |
| Diploma or Degree | 4 (4.1%) | 95.0 | 90.0-100% |
| Secondary education only | 81 (83.5%) | 87.1 | 84.2-90.0% |
| Sexual activity |  | SUS score | 95% CI |
| Sex in the past 12 months | 62 (63.9%) | 89.9 | 87.2-92.7% |
| No sex in the past 12 months | 30 (30.9%) | 83.9 | 78.6-89.3% |
| Sexual activity undisclosed | 5 (5.2%) | 83.5 | 69.8-97.2% |
| Self-reported HIV status |  | SUS score | 95% CI |
| HIV negative | 83 (85.6%) | 88.8 | 86.2-91.3% |
| HIV positive | 11 (11.3%) | 79.3 | 68.9-89.7% |
| Unknown HIV status | 3 (3.1%) | 90.0 | 79.7-100.3% |
| PrEP status |  | SUS score | 95% CI |
| On PrEP | 24 (28%) | 86.6 | 81.0-92.1% |
| Not on PrEP | 62 (72.1%) | 89.7 | 87.0-92.4% |

AGYWs rated the AI Companion as highly usable with a SUS score of 87.7 (SD 12.7). The SUS, a widely validated and commonly used measure of perceived usability, provides a benchmark score of 68, above which a system is generally considered usable, while scores above 80 are classified as “excellent” and associated with high user satisfaction and likelihood of continued use.^26–28^ Usability scores were consistently high across all participants, regardless of age, sexual activity, PrEP use, or HIV status. Differences of approximately 10% were observed in the high school category and between participants who were living with HIV and those with unknown HIV status. However, the unknown HIV status group was small (n = 3). See Table 1.

We also plotted the mean scores of the Likert scales of the six “Intentions to Engage in HIV-care” dimensions at pre-and-post intervention. These means and their confidence intervals are displayed in Figure 1. While the scores across most dimensions were higher post-interaction than at baseline, the overlap of confidence intervals indicates that there is no statistically significant difference (increase or decrease) across any of the dimensions. The point estimate for each dimension increased post-interaction, with the exception of dimension six (How likely are you to seek HIV treatment services if you test HIV positive?).

**Figure 1:**
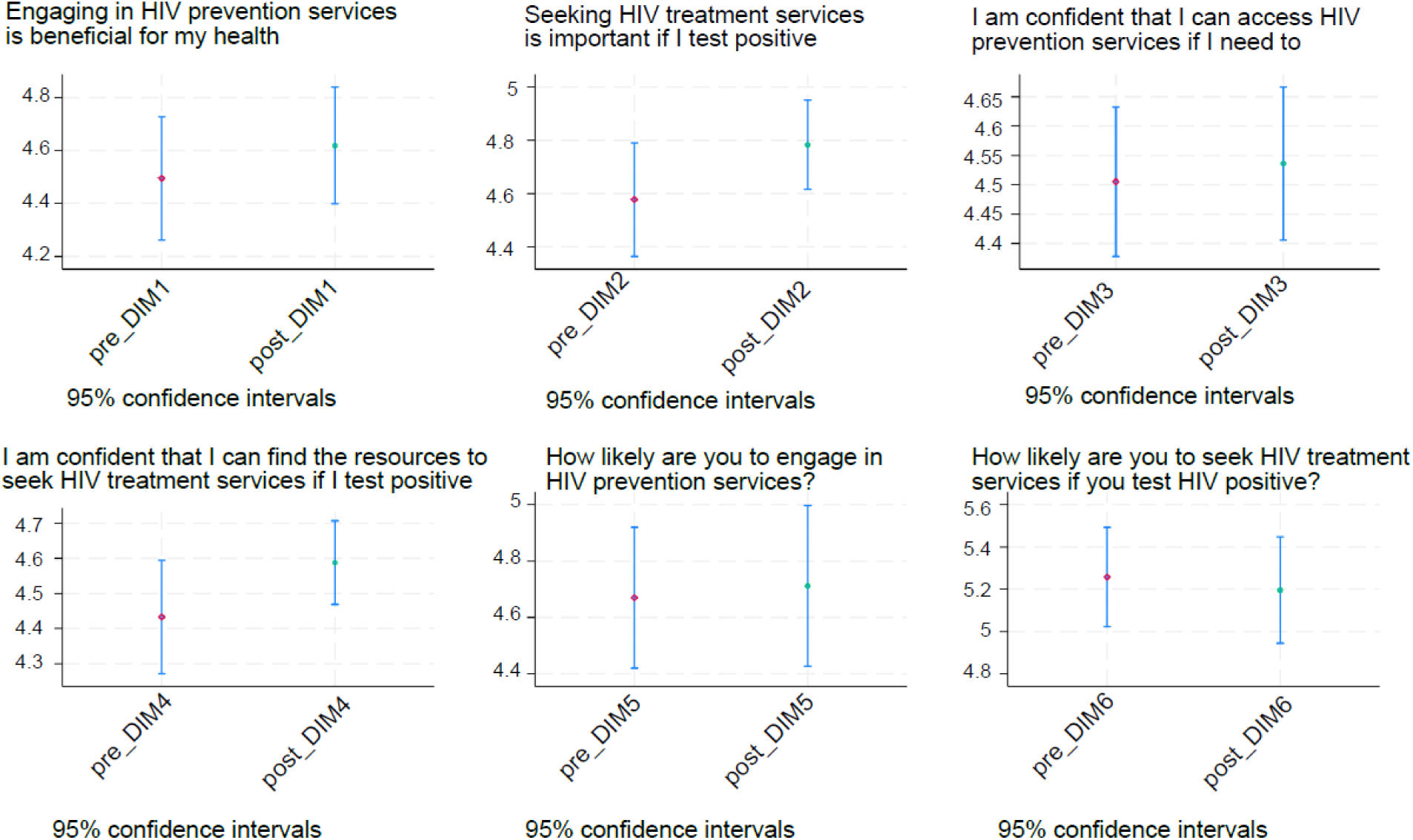
Mean score of the Likert scales of the “Intentions to Engage in HIV” dimensions pre-and-post the intervention.

### Healthcare providers

We enrolled 44 out of 50 healthcare providers from both public and private health sectors. Once we reached saturation of themes with 44 healthcare providers enrolled, we stopped further enrolments. The mean age for healthcare providers was 34 years (SD: 6.5), and about half were HTS (HIV testing services) counsellors. See Table 2 for baseline characteristics of healthcare providers.

**Table 2:** Characteristics of Healthcare Providers (n=44) enrolled in the study between November 2024 and May 2025.

|  |  | Healthcare providers<br>(N = 44) |
| --- | --- | --- |
| Age: Median (IQR) |  | 32.5 (30.5 - 37.5) |
| Highest tertiary qualification obtained: N (%) | Certificate<br>Diploma<br>Degree<br>Post grad qualification | 20 (45.5%)<br>9 (20.5%)<br>7 (15.9%)<br>8 (18.2%) |
|  |  | **Healthcare providers**<br>(N = 44) |
| Age: Median (IQR) |  | 32.5 (30.5 - 37.5) |
| Employment status: N (%) | Employed<br>Unemployed/full-time student | 44 (100%)<br>0 (0%) |
| Profession: N (%) | Professional nurse<br>Enrolled nurse<br>HTS Counsellor<br>Other* | 17 (38.6%)<br>1 (2.3%)<br>24 (54.6%)<br>2 (4.6%) |
| Duration in profession: N (%) | ≤ 1 year<br>1-3 years<br>≥ 4 years | 1 (2.3%)<br>11 (25%)<br>32 (72.7%) |
| Duration in profession: Mean (SD) | All | 2.7 (0.5) |

\* Profession Other - includes team leaders and operations support

Most healthcare providers rated the Clinical Portal ease-of-use, comprehension, and client support of the portal within the range of “positive” or “very positive”. However, 23% expressed concerns regarding workflow efficiency and their ability to manage additional client volume. See Figure 2 for a heat map showing provider responses to the post-survey questions on the Clinical Portal.

**Figure 2:**
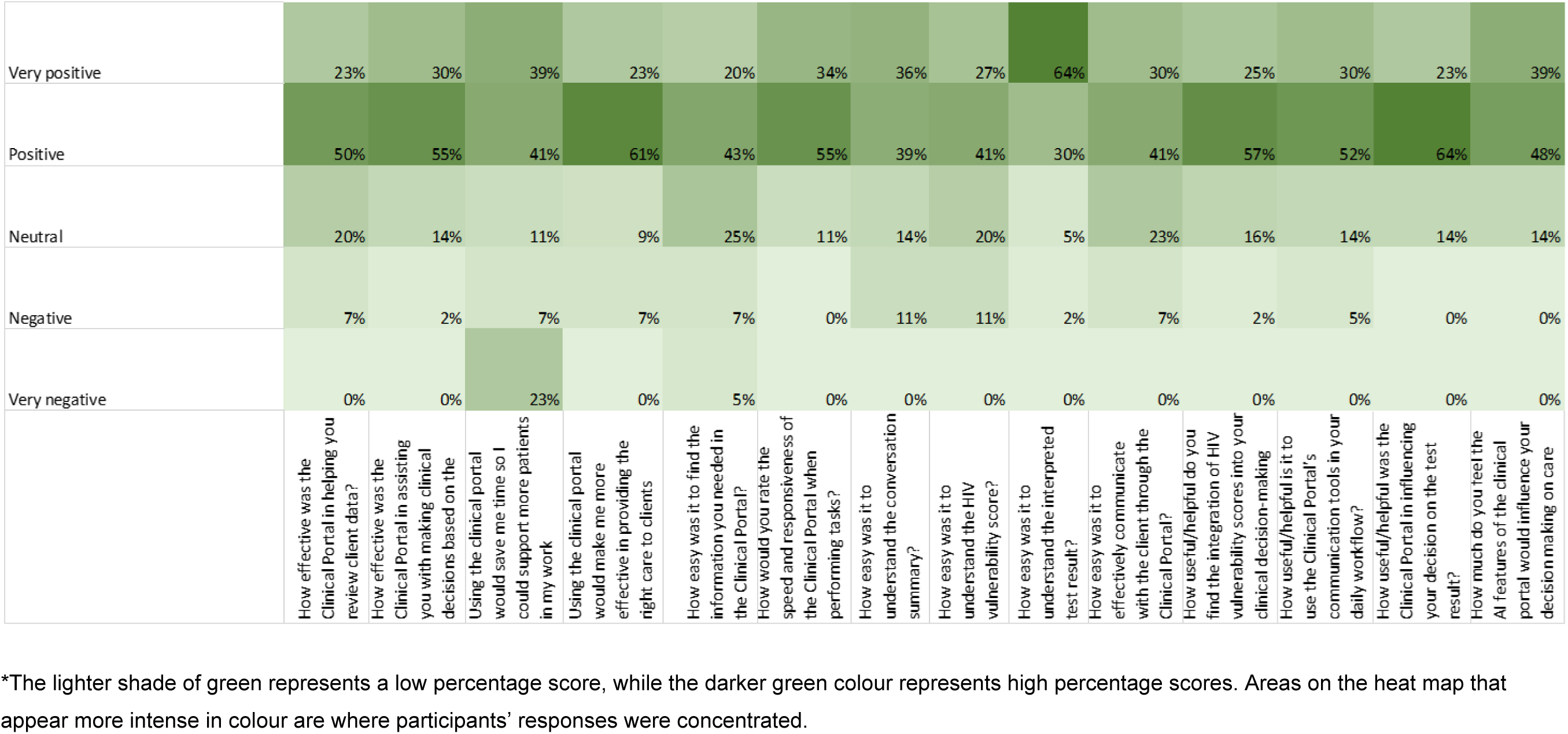
Heatmap showing provider perceptions of the Clinical Portal.

### Qualitative results

The findings are organised according to key principles of the User Centred Design (UCD) framework, which emphasises a deep understanding of users, their needs, behaviours, and preferences. UCD seeks to ensure that products and systems are intuitive and designed to support meaningful, user-friendly interactions. The themes below reflect how AGYW and healthcare providers engaged with and experienced the AI Companion and Clinical Portal, respectively.

### Adolescent girls and young women

We conducted a total of three focus group discussions (FGDs) with AGYW (N=20). Two FGDs were held with participants aged 18-24 years; one group included those who interacted with the AI Companion only, while the other included those who completed a simulated HIV self-testing (HIVST) activity. The third FGD included adolescents aged 16-17 years who participated in the simulated HIVST activity. Each FGD comprised between four and seven participants.

#### Understanding the AI Companion: benefits and comparisons to human interaction

Participants described the AI Companion as non-judgemental, confidential, and easy to engage with. Its tone and structure created a sense of comfort and emotional safety, enabling AGYW to discuss sensitive topics more freely.

> With AI, I was more comfortable with answering the questions they were asking because I do not know the AI… So, with the AI it’s like you are talking to a friend over [the] phone (AGYW FGD1, 01)

Participants acknowledged both the strengths and limitations of the AI Companion compared to human interaction. Most expressed a preference for the AI Companion due to its stigma-free and empathetic tone.

> Because if I were to tell a friend, they will be like, how could you do that, but the AI won’t tell me that. It will just guide me and give me advice and options or refer me to someone. (AGYW FGD1, 02).

Several participants contrasted the interactions with the AI Companion and prior experiences with healthcare providers. Negative attitudes from nurses made participants feel uncomfortable in the past, whereas the AI Companion’s neutrality allowed them flexibility to ask questions.

> I think other nurses have an attitude. You could be talking and explaining your story and the nurse starts giving you bad treatment. Whereas the AI Companion does not have an attitude. Once the nurse responds to me with a bad attitude, I will not feel comfortable. So, I prefer an AI because I am speaking with a person I don’t see. I am free to ask questions. (AGYW FGD2, 01).

#### Key Features: Personalisation, Privacy, and Information Delivery

AGYWs valued personalisation, privacy, and clarity of information delivered by the AI Companion. Participants emphasised that customising interactions allowed the AI to adapt to their individual needs.

> I chose customize, so you’re able to customize it and get it to explain it the way you understand like language barriers. When it gets to know you, it will explain things to you the way you would want to understand them. (AGYW FGD1, 3)

Privacy was another critical need, with participants sharing appreciation that the AI Companion did not request personal identifiers and that they were able to engage anonymously through the use of pseudonyms. This was perceived as essential for comfort and trust.

> Okay, it doesn’t ask about your personal information like ID, real name and it doesn’t ask more about your ID, your real name. They just want you to create your ideal name. (AGYW FGD1, 04)

Participants also valued the clarity of information delivery, noting that the AI Companion responded directly to the questions asked in a manner they could easily understand.

> The questions that you ask…get answered exactly the way you ask. It is not too different from the question. (AGYW FGD2, 02)

#### Credibility and limitations of the AI Companion

Participants generally valued the AI Companion for providing trustworthy, factual, and confidential responses, perceiving it as knowledgeable and helpful for accessing new and relevant health information.

> It really helped me because I thought that if you are pregnant and you have HIV…You tested positive, you use PrEP for an unborn child to not contact HIV, but I only found out today that you use ART. I did not know, so it did help. (AGYW FGD1, 05)

While many valued the conversational tone and emotional warmth the AI Companion conveyed, some noted potential limitations of digital companions related to trust in challenging situations. Some felt that the AI Companion may not be able to provide the level of emotional support required in crisis situations.

> “I felt like I was talking to a friend. I remember it asking me how I was feeling… and said Eish … and that made me feel like I’m talking to someone that I know.” (FGD1, 06)

> “I felt so comfortable because it didn’t have any problems, I didn’t get any problems when using it. Because it was so user friendly.” (AGYW FGD1, 04)

> “No, it is just an artificial tool. It does not have real feelings and emotions. No, I don’t [trust it]. Yeah, I do not trust AI.” (AGYW FGD1, 04)

> “…after testing, what if I killed myself, so with things like going to a doctor, we can actually get counselling after that. You can get proper treatment.” (AGYW FGD1, 01)

The AI Companion’s usability and convenience were highly valued. Participants highlighted features that made interactions easy, private, and practical, such as access to clinic locations and the ability to manage conversations while maintaining anonymity.

> “…it asked you if you want to delete your conversation or you want to keep it…” (AGYW FGD1, 04)

> “…it provides clinics near you where you can test for HIV.” (FGD1, 05) “…helped me to get that information that I needed faster.” (AGYW FGD1, 01)

### Healthcare providers

In addition, we conducted three FGDs (N=17) and two in-depth interviews (IDIs) with healthcare providers to explore their experiences and perspectives of using the Clinical Portal.

#### Understanding the portal and its effect on clinical practice

Providers described the Clinical Portal as a clinician-facing tool designed to provide visibility into the client’s AI Companion interactions that generates follow-up tasks to support nurse case management activities. Their initial understanding focused on its role in organizing client data, and a potentially valuable addition to their workflow, particularly for improving pre-consultation preparation and facilitating more personalised, informed interactions with clients.

> “basically that is a dashboard where [you] can respond to clients’ needs.” (Provider FGD2, 01)

> “So, for me the Clinical Portal gives you a summary of the chats from the chatbot. It also gives the task you’ve been assigned to assist the client. It can even manage clients’ cases.” (Provider IDI, 01)

Providers envisioned the portal as a tool to transform clinical processes by improving pre-visit preparation, with available history, and streamlining task management.

> “It will, because I will have the client’s background, and I will know how to deal with the client’s sensitive issues and on how to address them also.” (Provider IDI, 01)

> “For me I will say it will reduce the time I will spend with the client in terms of planning…” (Provider FGD2, 02)

#### Data quality and security considerations

Providers found the portal’s information, particularly chat summaries, to be a helpful starting point in making clinical decisions. Even in cases where they thought the summaries were not fully detailed, these still provided a solid basis for providers to engage clients and gather additional information as needed for comprehensive care.

> “…if that summary is clear and straight forward then we know what to do and we know what we face… a summary means everything is there at least we get some light to the tunnel to say this is what we have…we are dealing with this.” (Provider FGD2, 03)

> “They are helpful. But I think if you need more information you can chat with the client to get more information.”(Provider IDI, 01)

Providers were concerned that efforts to maintain client anonymity could complicate accurate record linking and increase the risk of duplicate client profiles on the Clinical Portal. Providers recommended that robust safeguards be implemented to ensure that each client’s information is accurately identified and consolidated.

> “I’m just thinking is there a way, I don’t know what can be used so that clients will not have duplicates – in a way? … That’s what I’m thinking that if someone returns and had to start over, will they also change [their] name again?” (Provider FGD3, 01)

#### User interaction and interface clarity

Some providers found the Clinical Portal straightforward to navigate, especially those more comfortable with digital systems. However, some experienced challenges understanding parts of the layout, locating certain features, and completing some of the tasks. Some interface elements were noted as occasionally unclear, these included locating the chat panel, discovering risk scores via hovering the mouse, and navigating the task list panel. Providers suggested that some portal elements should be named in a more actionable way, so users can easily understand their purpose.

> “…I think it’s easy if you’re used to a computer. Like knowing what you can get from where. Just because everything is there.” (FGD3, 02)

> “I would suggest a voice note where you can say something for 2 minutes… My thought is that the voice note makes things faster than typing.” (Provider FGD1, 01)

Overall, most providers found the Clinical Portal user-friendly, noting that the introductory videos helped them become familiar with its features and navigation. However, they indicated that additional training and hands-on experience would be needed to use the Clinical Portal confidently in routine practice.

## DISCUSSION

The use of artificial intelligence (AI) within healthcare is expanding rapidly; however, few studies have assessed the usability and acceptability of AI tools among potential users in the South African context. This study evaluated the usability, feasibility, and acceptability of the Self-Care From Anywhere (SCFA) toolkit, an AI-enabled digital health intervention designed to support HIV prevention, testing, and linkage to care among AGYW and healthcare providers in South Africa. The findings showed that both AGYW and healthcare providers found the SCFA toolkit, including the AI Companion and the Clinical Portal, to be usable, acceptable, and relevant to their needs. By triangulating quantitative and qualitative findings, we found high acceptability of both the AI Companion and the Clinical Portal, with qualitative insights suggesting that AGYW valued personalisation and privacy, while providers emphasised the Clinical Portal’s role in supporting case management.

Understanding users’ contexts, priorities, and care trajectories is critical in the design of digital health tools for diverse populations. The SCFA toolkit was perceived as useful and acceptable, and while overall usability (SUS score) was high, differences across certain sub-groups suggest that user characteristics; such as high school education and HIV status (HIV positive and unknown HIV status) may influence how participants engage with the AI Companion. In ongoing work we are seeing promising results of tailoring the tool for different user segments, ensuring that interventions appropriately address the distinct needs of different population groups.^30^

As a result of this study, additional iterations and adaptations of the SCFA toolkit include more content specific to treatment adherence, mental health, and social support for people living with HIV to enhance relevance and usability within this population.

Digital tools have the potential to support and improve intentions to engage in HTS when aligned to user needs. In this study, interaction with the SCFA toolkit was associated with an increase in self-reported intentions to engage in HIV prevention and care among AGYW. While these changes should be interpreted as proximal indicators rather than confirmed behavioural outcomes, they are consistent with prior evidence demonstrating that digital and mobile-based interventions can improve HIV-related knowledge, motivation, and behavioural intentions among adolescents and young adults.^31–32^ Features such as personalisation, confidentiality, and stigma-free AI Companion interactions; hallmarks of the SCFA toolkit have similarly been shown to enhance young people’s willingness to access HIV information and services when tools are perceived as empowering and non-judgmental.^33–34^

In the same vein, the overall positive reception of the Clinical Portal highlights the potential of digital health tools to support client management, task prioritisation, and more personalised provider-client interactions.^35–39^ One survey item assessing whether the portal could save time and enable providers to see more clients, elicited more mixed responses compared to other dimensions of usability and acceptability. This may reflect the complexity of provider workflows in resource-constrained settings, where improvements in information access and care quality do not always translate into immediate time savings or increased service capacity. Rather than indicating a limitation of the tool, this finding underscores the importance of understanding how digital systems are integrated into routine practice, including their effects on cognitive load, task sequencing, and perceptions of care quality. Future programme implementation should therefore examine real-world use of the Clinical Portal over time to better understand how efficiency gains may emerge as providers become more familiar with the system and as workflows adapt.

Applying behavioural insights to the design of digital health tools can support acceptability and integration of these services within a public health setting. The growing acceptance of AI-enabled tools among young people presents an important opportunity to strengthen HIV prevention and care pathways, particularly for populations that face barriers to traditional services. Studies across sub-Saharan Africa and globally have shown that adolescents are receptive to AI-powered and chatbot-based tools for sexual and reproductive health, mental health, and HIV support, largely because these tools offer privacy, urgency, and interactive engagement.^40–41^ Consistent with this literature, participants in this study described the AI Companion as convenient and accessible for discussing sensitive health topics. At the same time, sustaining engagement with AI-enabled interventions requires ongoing attention to trust, cultural relevance, and bias mitigation, particularly among marginalised youth who may experience digital exclusion or health system mistrust.^42–43^

This study had several limitations. First, as an exploratory study, the sample size was small and intended to provide descriptive insights rather than confirm statistically significant effects. The primary focus on usability and acceptability limits the generalisability of findings beyond the study context. Second, the study relied on self-reported data for sensitive measures such as HIV and PrEP status. While reports of HIV-positive status are likely accurate, HIV-negative or unknown status may be underreported, which may influence the interpretation of results concerning HIV and PrEP. Third, participants were drawn from existing service delivery programmes and within urban communities, which limits representation of harder-to-reach or digitally disconnected populations. The sample also likely included individuals already engaged with care, and findings may not fully reflect the experiences or needs of those less connected to healthcare services. Forth, while the AI Companion was delivered in English, South Africa’s linguistic diversity means that the tool may have been more accessible to participants comfortable communicating in English. Future multilingual adaptations may therefore be necessary to ensure equitable access and usability across different language groups. Fifth, for the Clinical Portal, we did not assess healthcare workers’ baseline familiarity with AI or prior exposure to digital health tools, which may have influenced usability perceptions. Finally, we did not provide formal training on the Clinical Portal, opting instead for a brief introduction to capture first-use impressions, which may differ from experiences following structured onboarding.

Despite these limitations, Audere has taken the findings from this study, adapted and developed improved iterations of this AI tool that are presently being evaluated through larger-scale implementation studies (e.g. Aimee) and operational programs such as Self-Cav.^44^ Further work is also needed to strengthen health economic evidence, including implementation costing and cost-effectiveness analyses, to inform investment decisions and sustainable scale-up.

## Conclusion

Leveraging artificial intelligence (AI) in digital health tools has significant potential to enhance client engagement, support healthcare providers, and improve health outcomes. This study demonstrated that the AI-powered Self-Care From Anywhere (SCFA) toolkit was both highly usable and acceptable among adolescent girls and young women (AGYW) as well as healthcare providers. However, the findings highlight the need for ongoing user-centred refinement to ensure that AI-driven tools remain relevant, and responsive to the diverse needs of users, whose experiences and care expectations may differ based on demographics, location, behaviours, HIV status, and more.

There is also a critical tension between client and provider priorities. AGYW valued the stigma-free, anonymous nature of the platform particularly for sensitive topics whereas healthcare providers emphasised the importance of client identification, comprehensive background information, and assured linkage to care to prevent duplication and ensure continuity within the health system. As AI-powered self-care tools, especially those leveraging large language models (LLMs) for confidential counselling become more embedded in care pathways, this mismatch between user preferences for anonymity and system-level requirements for identification may become increasingly pronounced. Future research should examine where, how, and at what stages identification becomes a necessary component of safe and effective care delivery, and how AI Companions and AI-enabled healthcare provider tools can be designed to maintain client confidentiality and autonomy while still supporting appropriate linkage, documentation, and continuity of care. Addressing this tension will be essential to realising the full promise of AI-driven self-care within equitable and responsive health systems.

## Ethics approval

The study was approved by ethics committees at the University of the Witwatersrand (ref: 240906) and Boston University (H-45383).

## Data Availability

All data produced in the present study are available upon reasonable request to the authors.

## Acknowledgements

We would like to thank Ms. Maaya Sundaram from the Gates Foundation for her guidance throughout this project. We are grateful to Shout-It-Now for their support in recruiting participants and to Ms. Busisiwe Sibiya for leading data collection. We would like to thank Phylaxis.ai for their development of the predictive risk model and partnership on the novel approach to collection of HIV acquisition vulnerability factors. We sincerely thank all participants for generously sharing their time and insights, which made this study possible.

## Data availability statement

Data are available on request to the corresponding author.

---

**Appendix 1:**
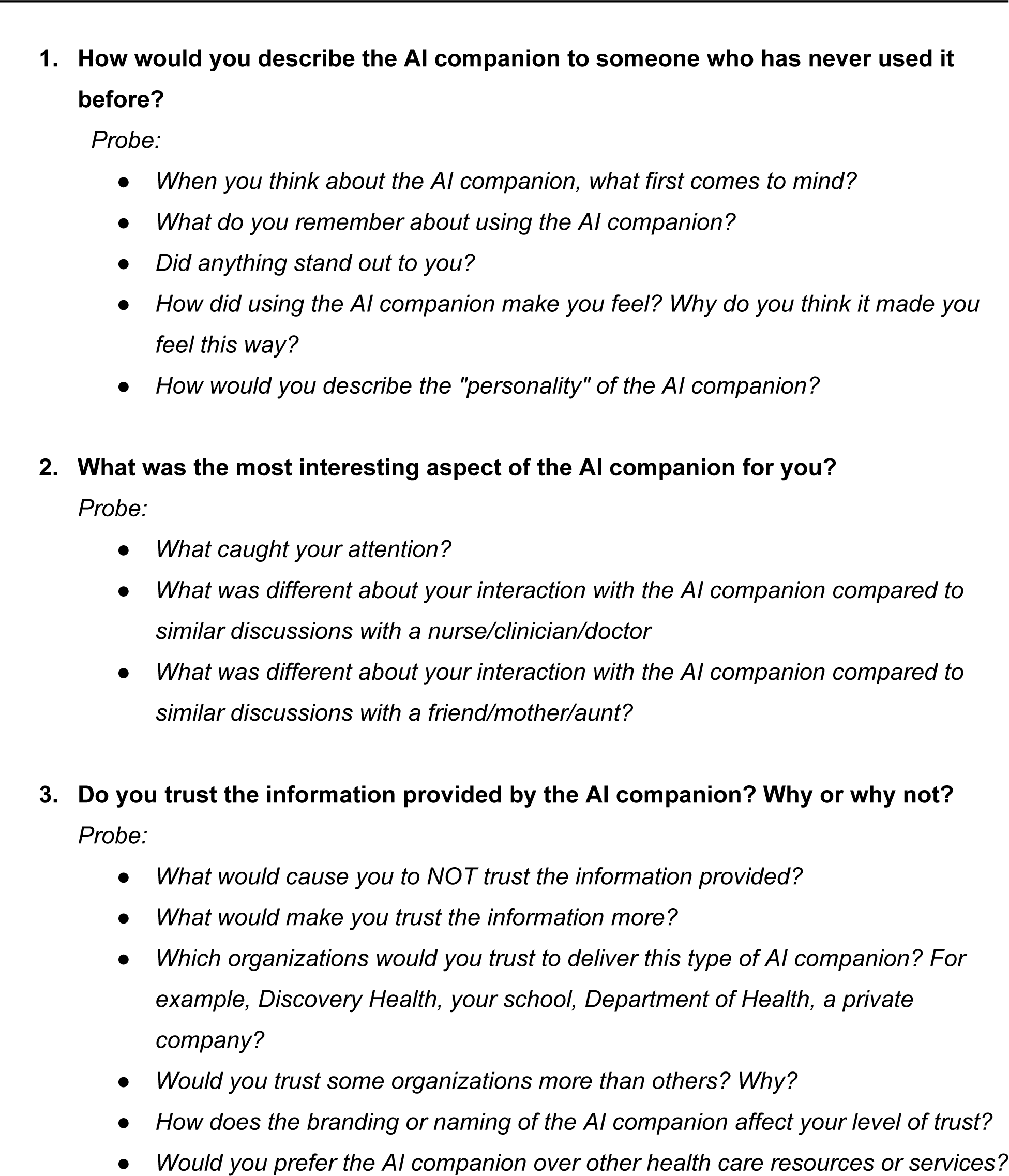
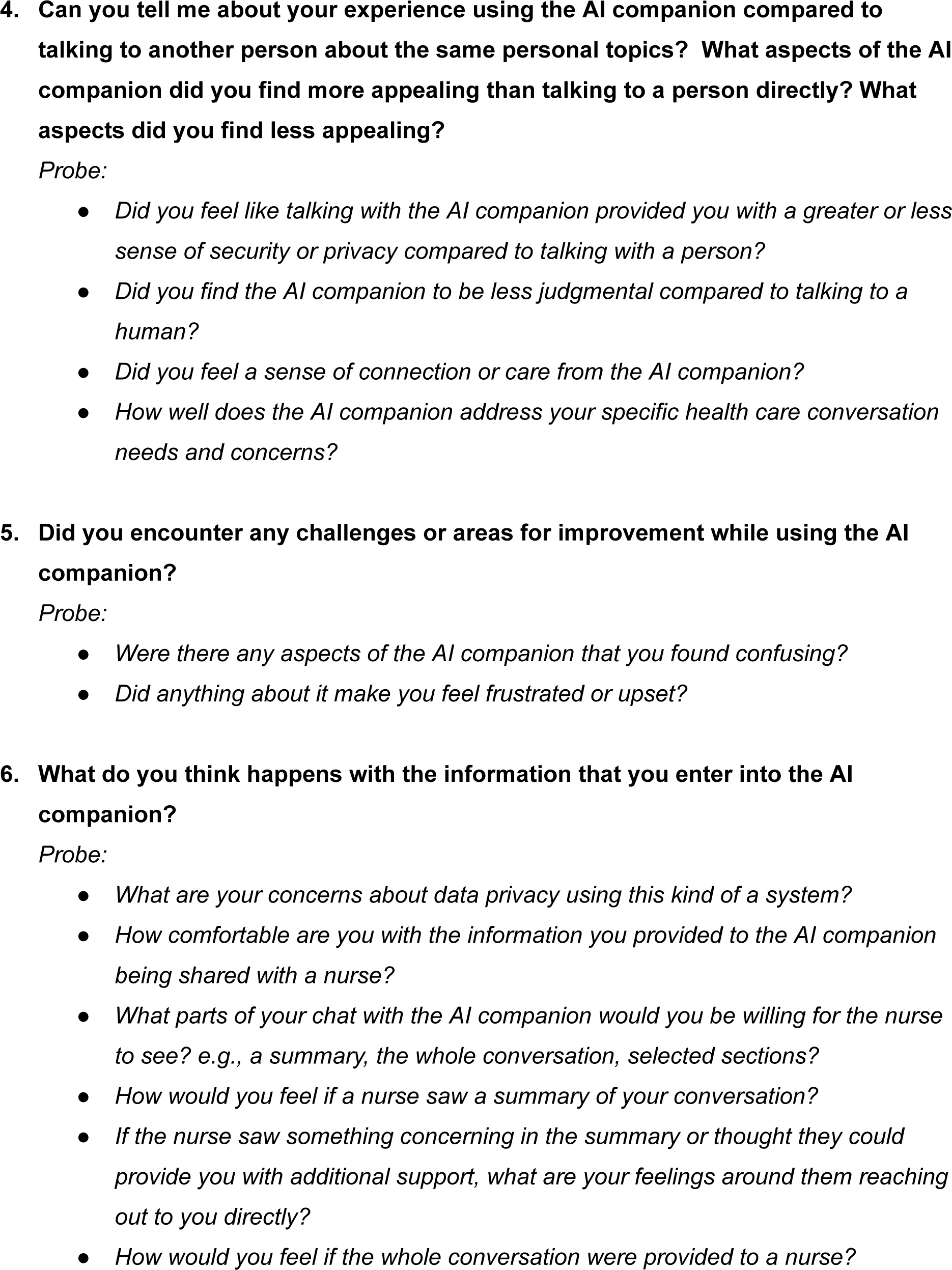
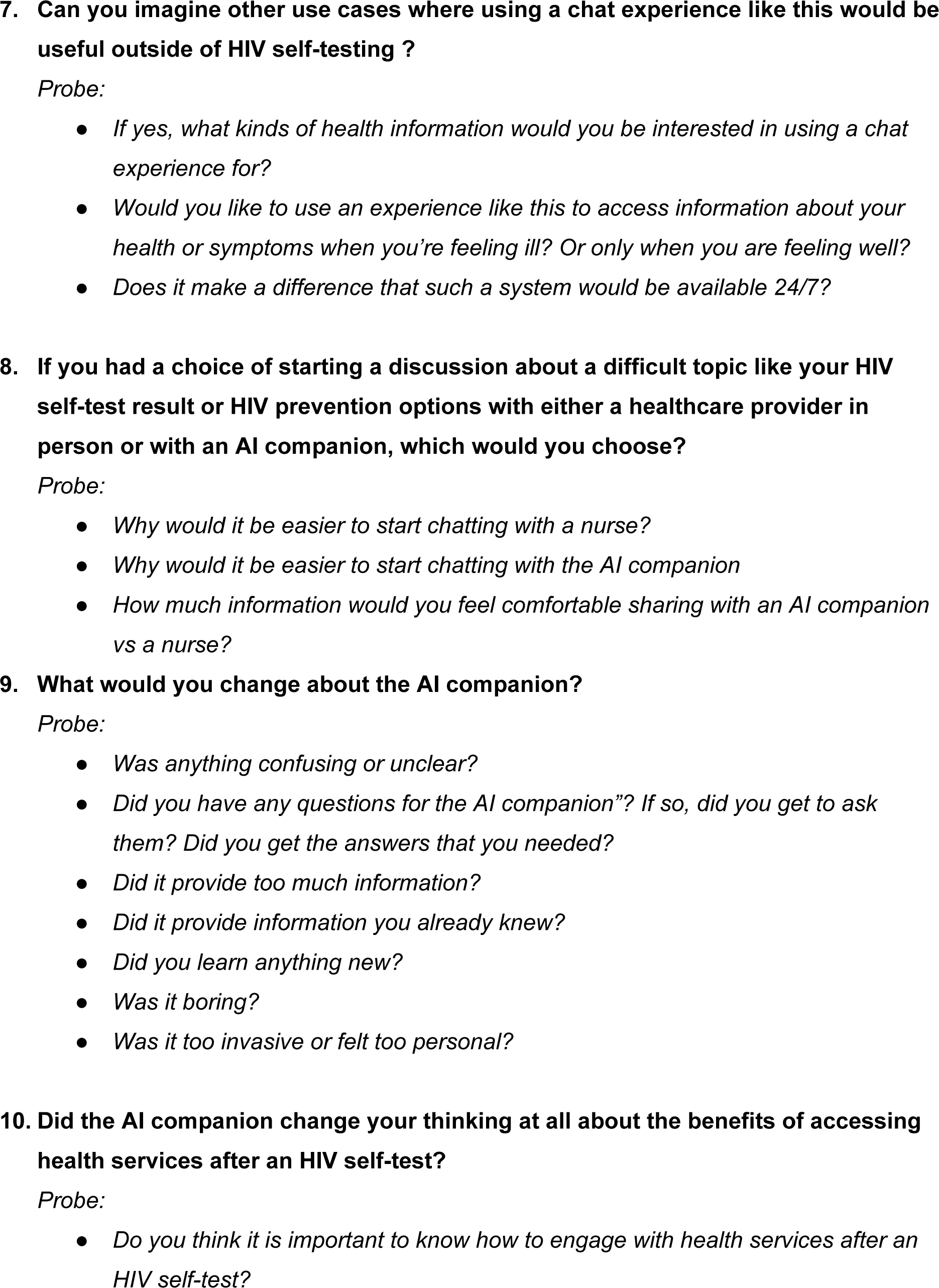
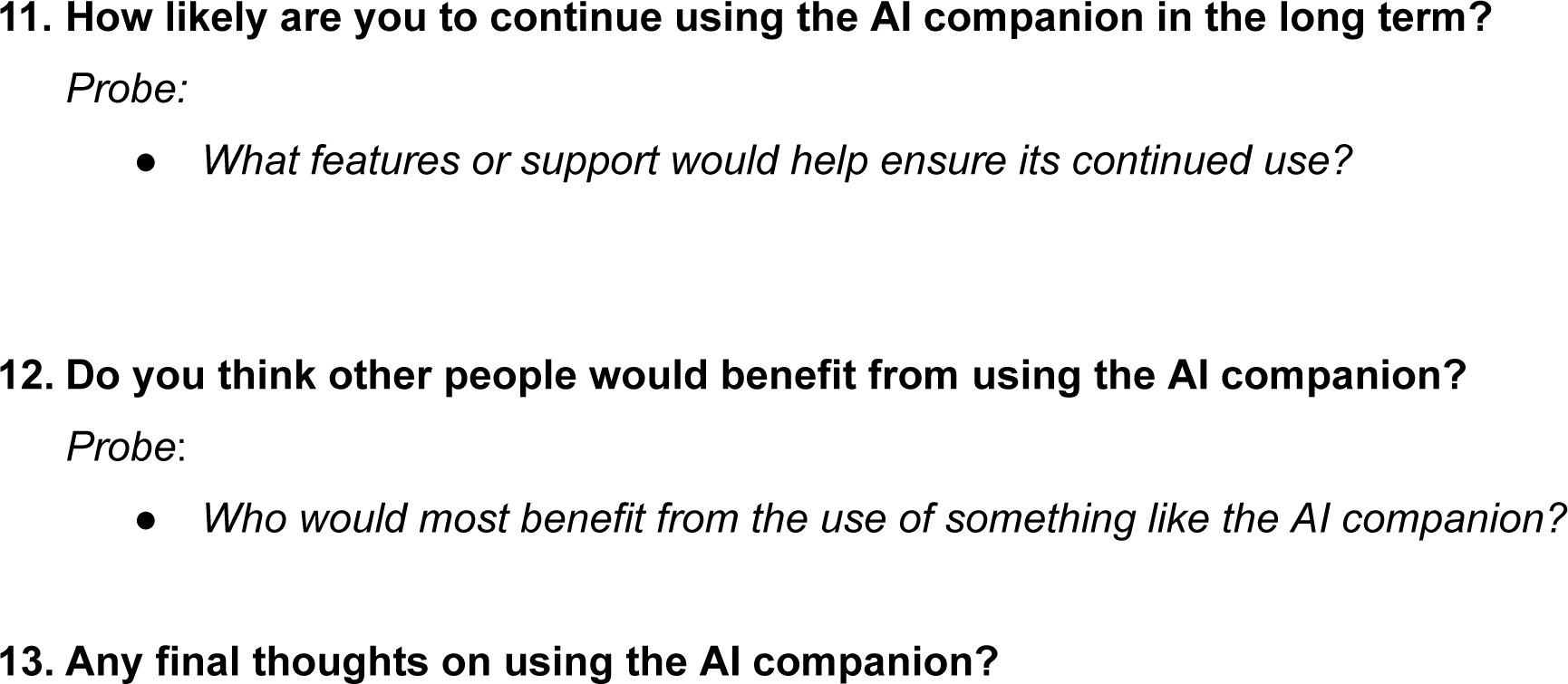
Focus group discussion guide for AGYW.

**Appendix 2:**
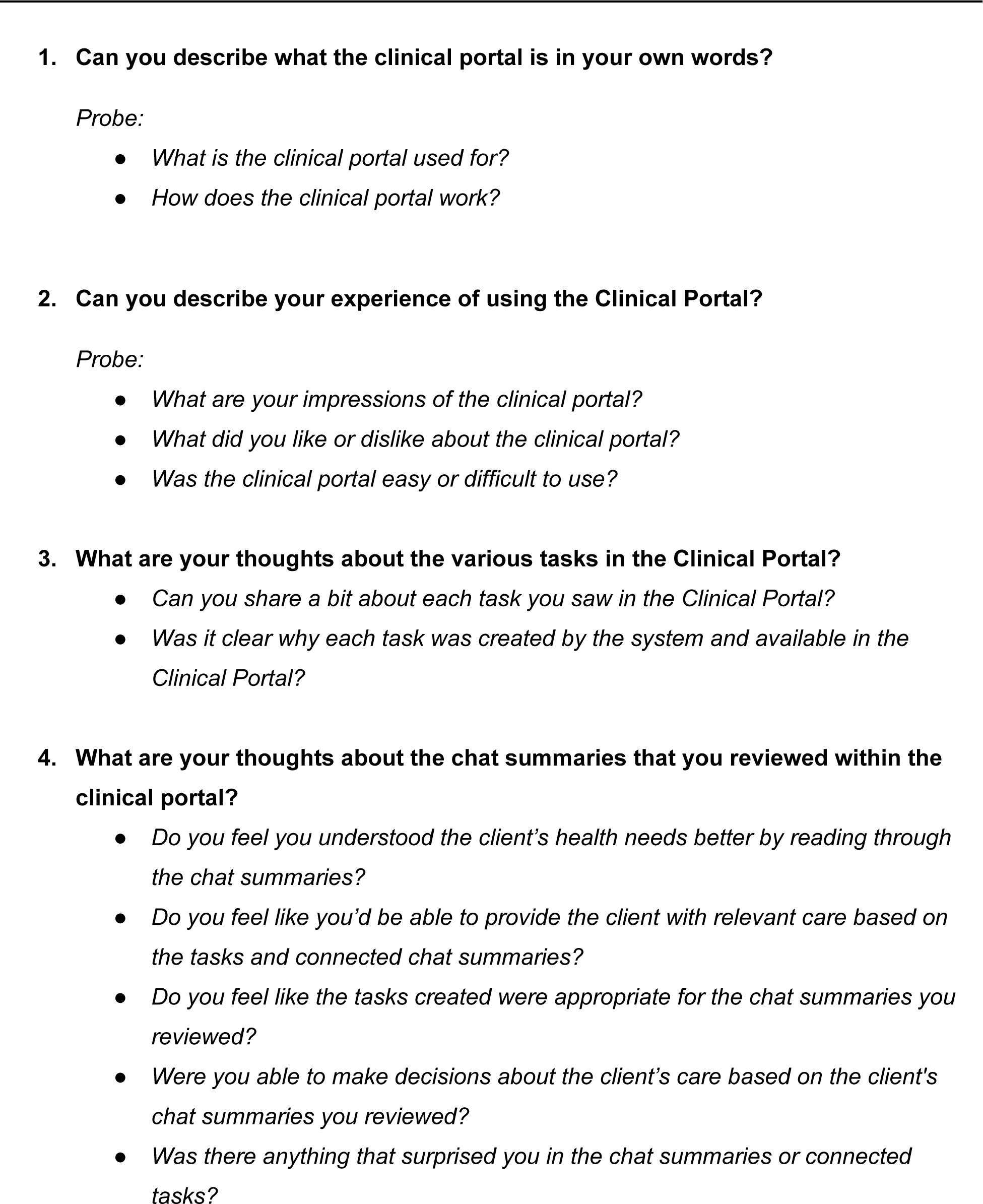
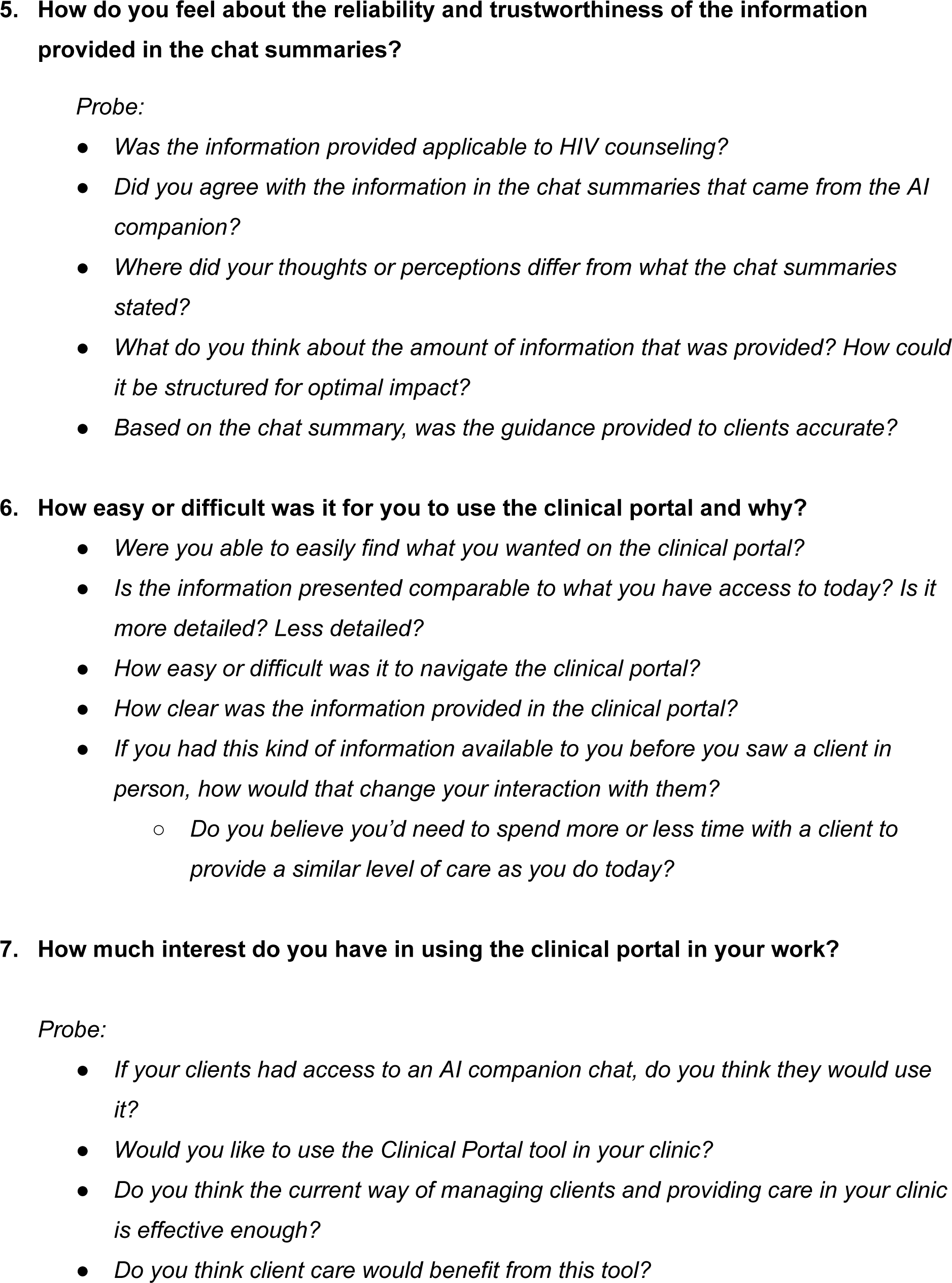
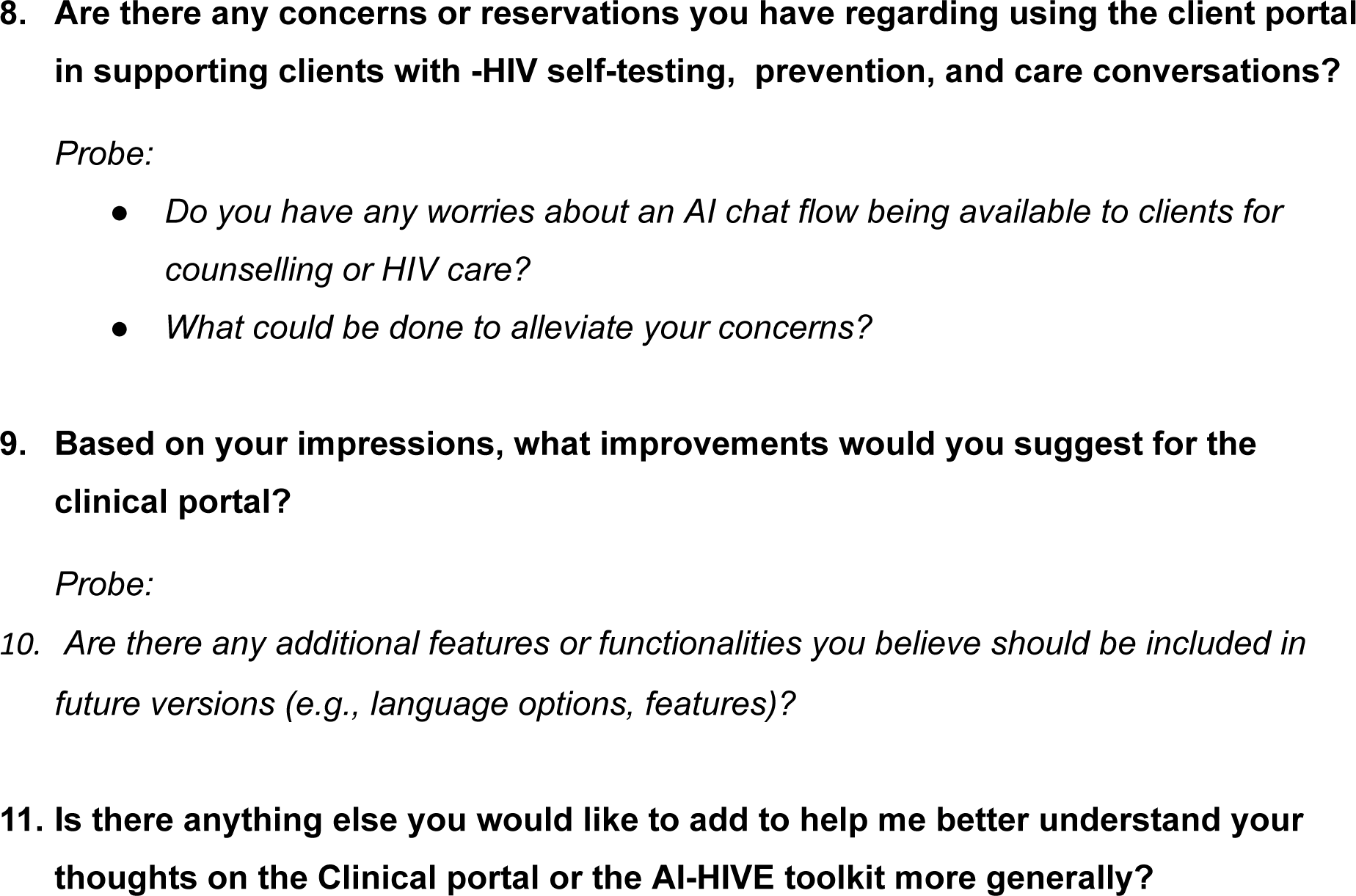
FGD & IDI guide for healthcare providers who engage with the Clinical Portal.

